# Hydroxychloroquine Safety Outcome within Approved Therapeutic Protocol for Covid-19 Outpatients in Saudi Arabia

**DOI:** 10.1101/2020.08.16.20175752

**Authors:** Abdulrhman Mohana, Tarek Sulaiman, Nagla Mahmoud, Mustafa Hassanein, Amel Alfaifi, Eissa Alenazi, Nashwa Radwan, Nasser AlKhalifah, Ehab Elkady, Abdullah Almohaizeie, Fouad AboGazalah, Khaled AbdulKareem, Fahad AlGhofaili, Hani Jokdar, Fahad Alrabiah

## Abstract

**Background:** Healthcare systems globally has been challenged following the COVID-19 pandemic, since late 2019. Multiple approaches and strategies have been performed to relieve the pressure and support existing healthcare systems. The Saudi Arabian Ministry of Health (MOH) launched an initiative to support the National Healthcare System. Since the 5^th^ of June 2020, 238 outpatient fever clinics were established across Saudi Arabia.

**Methods:** A cross-sectional study included 2,733 eligible patients subjected to MOH treatment protocol (hydroxychloroquine and zinc) and revisited the clinics within 3-7 days after treatment initiation. This study aimed to assess the safety outcome and reported adverse events from hydroxychloroquine use among suspected COVID-19 patients. The data was collected through an electronic link and cross-checked with that of the national database (Health Electronic Surveillance Network, HESN) and reports from the MOH Morbidity and Mortality (M&M) Committee.

**Results:** Majority of the cases were males (70.4%). Upon reassessing the studied participants within 3-7 days, 240 patients (8.8%) discontinued the treatment protocol because of the development of side effects (4.1%) and for non-clinical reasons in the remaining (4.7%). Medication side effects overall were reported among (6.7%) of all studied participants, including mainly cardiovascular adverse events (2.5%), followed by gastrointestinal (GI) symptoms (2.4%). No Intensive Care Unit (ICU) admission or death were reported among these patients.

**Conclusion:** In our study, results show that the use of hydroxychloroquine for COVID-19 patients in mild to moderate cases in an outpatient setting, within the protocol recommendation and inclusion/exclusion criteria, is safe, highly tolerable, and with minimum side effects.

## BACKGROUND

COVID-19 has rapidly emerged as a pandemic infection that has caused significant morbidity and mortality worldwide. Global healthcare systems had faced multiple challenges varying from the high number of visitors to lack of approved therapeutically options, since the outbreak of SARS-Cov-2 late 2019. Extensive efforts have been made to explore effective therapeutics against the virus globally (1). The World Health Organisation (WHO) states that there is no current evidence to recommend any specific anti-COVID-19 treatment for patients with confirmed COVID-19 (2).

Hydroxychloroquine has antiviral effects in vitro, and, in association with azithromycin, was suggested to decrease SARS-CoV-2 viral load in a small, non-randomised study (3,4). However, observational studies have suggested no beneficial effect of chloroquine or hydroxychloroquine in hospitalised patients with Covid-19 (5,6).

The Saudi Arabian Ministry of Health (MOH) has launched an initiative to support the National Healthcare System. Since the 5th of June 2020, 238 outpatient fever clinics were established across the Kingdom, under the direct supervision of the Primary Healthcare Deputyship at the MOH. The target population for these clinics was suspected COVID-19 patients showing mild to moderate symptoms, and the implemented protocol was based on hydroxychloroquine + zinc sulphate as the recommended treatment of choice.

Given the hydroxychloroquine safety profile, multiple exclusion criteria, and several recommendations for patient safety were implemented in the protocol. Baseline ECG and electrolyte results were requested before initiation to overcome and follow-up the risk of cardiac arrhythmias leading to sudden death (if happened). On the other hand, high-risk patients for developing side effects, especially cardiac, were excluded from the hydroxychloroquine use and advised to follow standard care.

Evidence regarding the potential therapy of hydroxychloroquine, whether given alone or in combination with zinc sulphate, for COVID-19 patients, is not clear and limited. Therefore, this study aimed to assess the safety outcomes and adverse events among COVID-19 patients attending outpatient fever clinics and subjected to the MOH approved treatment protocol within 3-7 days in Saudi Arabia. This publication is part of a national project to assess the safety outcomes from the national fever clinic initiative in Saudi Arabia.

## OBJECTIVES

To determine the reported adverse events among suspected COVID-19 patients subjected to the MOH approved treatment protocol and assessed within 3-7 days, including the development of treatment-related side effects, medication tolerability, hospital admission, ICU admission, and death.

## METHODS

### Study design and setting

A cross-section study was conducted from the 5^th^ of June to the 7^th^ of July 2020, in (238) outpatient fever clinics in Saudi Arabia.

### Study participants

The sample size included 2,733 eligible COVID-19 suspected patients subjected to MOH treatment protocol before receiving PCR results. Out of 60,738 (37.9%) consecutive COVID-19 suspected patients who attended clinics during the study period, 23,043 (37.9%) eligible patients were given the approved treatment protocol. Out of these, 2,733 patients who revisited the clinics within 3-7 days and fulfilled the inclusion criteria were included in the sample and reassessed for adverse events and safety outcome of the approved treatment protocol.

The study included patients aged 19 years and above, presenting with subjective fever (> 38 °C), and with one or more COVID-19 symptoms including; runny nose, sore throat, cough, diarrhoea, shortness of breath, headache, and myalgia and those who revisited the clinics within 3-7 days. The study excluded morbidly obese patients, pregnant and lactating females, those with G6PD deficiency, and patients with known cardiac-related issues.

### Study tools

Data from patient health records was entered into pre-designed advanced online forms by data entry officers at the local Medical Affairs. The data collected through an electronic link and cross-checked with the health electronic surveillance network database and reports from the MOH Morbidity and Mortality (M&M) Committee. All collected data were sent to the District Medical Affairs for follow-up and investigation to determine the safety outcome of the treatment protocol during the first three days, including medication discontinuation, reported adverse drug reactions in terms of GI symptoms and ECG abnormalities, complete recovery of COVID-19 symptoms, hospitalisation, ICU admission, and death.

A standardised prescription form was written and distributed within the written treatment protocol. The prescription list hydroxychloroquine for five days duration, 400mg twice for day one, followed by 200mg twice daily for the remaining duration. The prescription also includes zinc sulphate 60mg orally once daily for five days, paracetamol and antihistamine. The prescription did not include azithromycin into it.

### Statistical analysis

The outcome variables included reported adverse events and safety outcome of the treatment protocol used, including the percentages of patients hospitalised, admitted to ICU, death, medication discontinuation, and developed side effects, e.g., ECG changes, GI symptoms. All data were analysed using SPSS^®^ version 21.

### Ethical consideration

This research has met, approved, and been followed closely by the MOH Institutional Review Board (IRB). Informed consent was taken, by the treating physician, from the participants after explanation of the study. Those who refused to participate in the study were excluded and continued with stander care. All collected information will be kept confidential and will not be used for other purposes than the study. Hydroxychloroquine therapy was discontinued at any time in patients who reported any possible medication-related adverse events. Unstable patients at presentation or those who showed clinical progression/deterioration at day three were referred to the hospital and continuously followed up by the research team for outcomes.

## RESULTS

Table (1) presents the socio-demographic characteristics among studied participants. Out of 2,733 patients, 56.89% were PCR positive, and the remaining were either PCR negative or their results were not obtained. Most of them were males (69.4% and 71.8% of both groups respectively). More than one third (36.3%) of the studied patients were aged 31-40 years, and (20.6%) were 41-50 years old. When the patients were reassessed within 3-7 days, a total of 183 patients (6.7%) had developed medication-related side effects, and 240 patients(8.8%) had discontinued the treatment protocol.

**Table (1):** Socio-demographic characteristics among studied both PCR positive and negative participants (n = 2,733).

|  |  | PCR positive<br>(n=1555) |  | PCR Negative /NA<br>(n=1178) |  | Total<br>(n=2733) |  |
| --- | --- | --- | --- | --- | --- | --- | --- |
|  |  | Frequency | % | Frequency | % | Frequency | % |
| Gender | Male | 1079 | 69.4 | 846 | 71.8 | 1925 | 70.4 |
|  | Female | 476 | 30.6 | 332 | 28.2 | 808 | 29.6 |
| Age | 19-30 | 447 | 28.7 | 372 | 31.6 | 819 | 30.0 |
|  | 31-40 | 567 | 36.5 | 426 | 36.2 | 993 | 36.3 |
|  | 41-50 | 320 | 20.6 | 243 | 20.6 | 563 | 20.6 |
|  | 51-64 | 215 | 13.8 | 135 | 11.5 | 350 | 12.8 |
|  | 65+ | 6 | 0.4 | 2 | 0.2 | 8 | 0.3 |
| Discontinuation of treatment | Yes | 143 | 9.2 | 97 | 8.2 | 240 | 8.8 |
|  | No | 1412 | 90.8 | 1081 | 91.8 | 2493 | 91.2 |
| Medication side effects | Yes | 100 | 6.4 | 83 | 7.0 | 183 | 6.7 |
|  | No | 1455 | 93.6 | 1095 | 93.0 | 2550 | 93.3 |

Out of 240 patients who discontinued the treatment protocol, one hundred and twelve patients (46.7%, 4.1% of total) developed medication side effects, and the remaining 128 patients (53.3%, 4.6% of total) discontinued the treatment protocol for non-clinical reasons not related to medication side effects (Table 2).

**Table (2):** The frequency of developed medication side effects among patients who discontinued the treatment protocol (n = 240)

|  |  | Cases discontinued treatment (n=240) |  |
| --- | --- | --- | --- |
|  |  | Frequency | % |
| Medication side effects | Medication side effects | 112 | 46.7 |
|  | No medication side effects | 128 | 53.3 |
|  | **Total** | **240** | **100** |

Out of 183 patients (6.7% of total) who developed medication side effects, one hundred and twelve (61.2%, 4.1% of total) discontinued the treatment protocol, and the remaining 71 patients (38.8%, 2.6% of total) continued the treatment irrespective of side effects (Table 3).

**Table (3):** Discontinuation of the approved treatment protocol among patients who developed side effects (n = 183).

|  |  | Cases reported medication side effects (n=183) |  |
| --- | --- | --- | --- |
|  |  | No. | % |
| Cases discontinued treatment | Discontinued treatment | 112 | 61.2 |
|  | Continued treatment | 71 | 38.8 |
|  | **Total** | **183** | **100** |

Table (4) explains the frequency of the reported medication side effects among studied participants. The most common reported side effect was cardiovascular symptoms (69 patients, 37.7%, 2.5% of total), including palpitation, chest pain, ECG changes (from baseline; QTc prolongation, arrhythmias), followed by GI symptoms, including nausea, vomiting, abdominal pain, diarrhoea.

**Table (4):** The frequency of reported side effects among studied PCR positive cases (n = 183).

|  | PCR positive cases (n=100) |  | Total (n=183) |  |
| --- | --- | --- | --- | --- |
|  | No. | % | No. | % |
| Cardio-vascular symptoms (palpitation, chest pain, ECG changes*) | 45 | 45.0 | 69 | 37.7 |
| GI symptoms (Nausea, vomiting, abdominal pain & diarrhoea) | 27 | 27.0 | 65 | 35.5 |
| Cardio-vascular symptoms + GI symptoms | 3 | 3.0 | 5 | 2.7 |
| Itching/medication hypersensitivity | 2 | 2.0 | 6 | 3.3 |
| Visual complication | 3 | 3.0 | 3 | 1.6 |
| Dizziness | 0 | - | 2 | 1.1 |
| Electrolyte abnormalities compared to baseline | 1 | 1.0 | 2 | 1.1 |
| Others (less commonly reported) | 19 | 19.0 | 31 | 16.9 |
| **Total** | **100** | **100.0** | **183** | **100.0** |

\* ECG changes: changes in ECG reading compared to baseline, QTc prolongation, arrhythmias

In all reported cases with medication-related side effects, no ICU admission or death was reported in patients who received hydroxychloroquine. Final adverse events reported cases would be revised with the final ongoing project analysis.

## DISCUSSION

The study included 2,733 eligible patients who were subjected to MOH treatment protocol and revisited the clinics within 3-7 days after initiation of therapy. Most of them were males (70.4%) in the age group 19-30 and 31-40 years (30.0% and 36.3% respectively). On reassessment of the studied participants within 3-7 days, 240 patients (8.8%) discontinued the treatment protocol because of the development of side effects (46.7%) and for other non-clinical reasons not related to medication side effects in the remaining (53.7%).

Hooks et al. studied the effects of hydroxychloroquine treatment on QT interval among 734 patients (mean age = 64.0 ± 10.9 years), of whom (90%) were men. They recorded an increase in mean QTc from (424.4 ± 29.7ms) to (432.0 ± 32.3ms) (P < 0.0001) during hydroxychloroquine treatment and found that Chronic Kidney Disease (CKD), history of atrial fibrillation (AF), and heart failure were independent risk factors for prolonged QTc (7). A more frequent prolongation of the corrected QT interval among patients receiving hydroxychloroquine, alone or with azithromycin, than in those who were not receiving either agent was also documented by Cavalcanti et al. and Lagier et al. (among 0.67% of patients) (8–9).

The current study recorded medication side effects among (6.7%) of all studied participants, including mainly cardiovascular symptoms (37.7%, 2.5% of total), e.g., palpitation, chest pain, ECG changes followed by GI symptoms, e.g., nausea, vomiting, abdominal pain, diarrhoea. Among those developed side effects, (61.2%, 4.1% of total) discontinued the treatment protocol. Although these symptoms were reported by patients during treatment course, we can not exclude the effect of COVID-19 overall symptoms to overlap patients reported adverse effects.

A higher percentage of adverse events was reported by Tang et al. and Lofgren et al. among hydroxychloroquine recipients’ patients (30%) compared to hydroxychloroquine non-recipient patients (9%), with patients reporting serious adverse events, the most common being diarrhoea, reported in 7/70 (10%) patients (10, 11). Similarly, Lofgren et al. reported a higher frequency of side effects (84%) and medication side effects (27%) among hydroxychloroquine recipients mainly; upset stomach (25% with daily, 18% with twice vs 23% for daily, 16% twice weekly for placebo), nausea (12% weekly, vs 6% for placebo) followed by diarrhoea, vomiting, or abdominal pain (11). This finding was inconsistent with Satlini et al., who found that patients with incident vomiting or diarrhoea were rare (12).

Other adverse events were recorded by Eljaalya et al. who investigated the pooled adverse effects of hydroxychloroquine among nine randomised trials in 916 patients and found that hydroxychloroquine caused significantly more skin pigmentation than placebo (Peto OR, 4.64; 95% CI, 1.13 to 19.00; *P=0.033*; I^2^ = 0%) (13). While they found other adverse events were not statistically significant; rash (Peto OR, 1.11; 95% CI, 0.3 to 3.77; *P=0.03*; I^2^ = 0%); gastrointestinal adverse events (Peto OR, 1.43; 95% CI, 0.55 to 3.72; *P=0.46*; I^2^ = 15.17%); headache (Peto OR, 1.94; 95% CI, 0.65 to 5.78; *P=0.23*; I^2^ = 9.99%); dizziness (Peto OR, 1.32; 95% CI, 0.49 to 3.52; *P=0.58*; I^2^ = 0%); fatigue (Peto OR, 2.13; 95% CI, 0.76 to 5.98; *P=0.15*; I^2^ = 0%); and visual adverse events (Peto OR, 1.61; 95% CI, 0.76 to 3.41; *P=0.22*; I^2^ = 0%). Inconsistent with our study, cardiac toxicity was not reported.

The current study recorded no ICU admission or death among studied participants. While Lagier et al. reported fatality rate cases were (0.9%) but they noticed a decreased risk of ICU admission and death (Hazard ratio (HR) 0.18 0.11 0.27), decreased risk of hospitalisation ≥ ten days (odds ratios 95% CI 0.38 0.27-0.54) among these patients treated with hydroxychloroquine-azithromycin.

The present study recorded (8.8%) discontinuation of hydroxychloroquine among studied participants due to the development of side effects (46.7%, 4.1% of total) and non-clinical reasons in the remaining (53.3%, 4.6% of total), which was consistent with Satlini et al. who found that eighty-nine percent of their patients completed the hydroxychloroquine course. Only three patients discontinued therapy because of QT prolongation (12).

## CONCLUSION

Overall, results show that hydroxychloroquine was tolerable within our patients and with a very minimum reported side effect. In addition, mortality and hospitalisation directly related to hydroxychloroquine treatment were not reported. Thus, our result can assure that the use of hydroxychloroquine for COVID-19 patients in mild to moderate cases in the outpatient setting, within the protocol recommendation and inclusion/exclusion criteria, is safe, highly tolerable, and with minimum side effects.

Due to the large number of patients who visited the clinic and lack of follow up visits within seven days, variables of safety measures were not obtained in this population. Therefore, the data regarding hospitalisation and death was cross-matched with that of the national M&M Committee, and regional Medical affairs across Saudi Arabia.

## Data Availability

All data are available upon request

